# “The Association of the IL-1RA (rs315952) Cytokine Gene Polymorphism and Susceptibility to Systemic Lupus Erythematosus: A Meta-analysis”

**DOI:** 10.1101/2024.11.16.24317426

**Authors:** Jan Mikkos L. Carlos, Mary Angelei T. Gabriel, Lark Joshua Gatbunton

## Abstract

Through a general retrospective analysis of the IL-1RAmspa11,100 (rs315952) cytokine gene polymorphism’s genotypes and alleles, the T/T genotype, along with the individual T allele, shows significant associations with the risk of Systemic Lupus Erythematosus (SLE). Meanwhile, the C/C and C/T genotypes, as well as the individual C allele, exhibited no significant associations. These results imply that having a homozygous genotype for the T allele increases the likelihood of SLE occurrence, while having a single C allele may induce a protective effect against the disease. To obtain these results, the researchers employed the analysis of six selected case-control and cross-sectional studies on healthy individuals SLE patients diagnosed through PCR tests. Forest plot construction was done to assess potential overlapping confidence intervals among the qualified studies. To identify the degree of heterogeneity among the papers, the odds ratio and the total confidence intervals, with the confidence interval set at 95%, were acquired through fixed and random effects models. Genotype and allele analyses with I2 statistics higher than 40% led to the funnel plot construction for outlier detection. Currently, the underlying processes that led to these genes’ and alleles’ varying outcomes to SLE risk remain elusive. Meanwhile, the subjects’ IL-1RA haplotypes may undermine these findings, along with the need for environmental consideration and the potential alteration of the gene and its corresponding alleles due to epigenetics. However, these findings still remain a possible starting point or reference for future studies.

## I. INTRODUCTION

One of the statements of the Centers for Disease Control and Prevention (CDC) highlight systemic lupus erythematosus (SLE) as a systemic autoimmune disease that predominantly occurs among women, and non-Caucasians [1], leaving approximately 200,000 cases of this autoimmune disease in U.S.A. alone [2]. The incidence of acquiring SLE may affect multiple organs simultaneously while severely impacting one’s central nervous system [3]. Among the various manifestations SLE can exhibit, hematological complications such as neutropenia, which is marked as a deficiency of the most abundant WBC, are conditions that are common among SLE patients. The exact pathogenesis of this disease remains elusive, raising questions as to what environmental and genetic factors are highly involved towards developing the disease [4]. However, it had been established that immune responses are activated due to the interactions that arose between the individuals’ genes and the external environment, leading to an overregulation of the plasma B cells, and as well as the effector cells that produce cytokines which may prompt for a cytokine storm, with some of these variants from the interleukins, interferons, chemokines, and other mediators as possible candidate genes for developing SLE [<u>5</u>; <u>6</u>]. The plasma B cells (B cells) are cells that are not synthesized in the liver, but rather, they are cells that produce antibodies by differentiating into mature B cells that play a key role in adaptive immunity as they are situated outside the bloodstream [7]. In terms of immunology, the CD4+ cells assist in the development of the B cells into plasma cells that activate antigen specific CD4 and T cells to aid in the release of cytokines which are potent to modify macrophages that repel foreign invaders [8]. The mechanisms behind these whenever overregulated may suggest SLE, such that the B cells are activated, in turn, activating autoreactivity even when these cells are not needed [9]. Other cells such as effector cells essential for adaptive immunity can also permit the emergence of SLE. These cells like CD8+ effector cells which is a type of T lymphocyte are prominent in directly killing the cells by releasing cytokines [10]. These effectors, when rendered dysfunctional, allow for the pathogenesis of SLE as a result of genetic defects and inflammatory cytokines that produce a double negative t-cell [11]. A double negative t cell is problematic as gamma-delta t cell receptors are yielded, but not the CD4+ helper t cells that aid in adaptive immunity [12]. Therefore, both effector cells and B cells are cells that play a role in cytokine production, such that the selected cytokine gene variants may induce SLE in a patient.

Cytokines are soluble proteins that are secreted by macrophages which are prominent at mediating cell reactions; therefore, these imply that some cytokines are essential at signaling the nervous system in times of diseases and tumors, while some cytokines may also become inflammatory cytokines like the TnF-α due to their capacity to allow other cell reactions that can pave the way for rheumatoid arthritis [13]. Just like any other protein, cytokines are in dire need of receptors to function. One of the types of cytokines enlist the interleukin which are cytokines that permit communication between WBCs to permit an efficient immune response [14]. One of the most important types of interleukin is the interleukin-1 (IL-1) which is a family of ligands and receptors known to be produced by macrophages in large amounts for defensive reactions [15]. For the current meta-analysis, the IL-1R was emphasized.

The interleukin-1 receptor (IL-1R) is a protein that aids in transmitting signals within cells in response to the inflammatory potent and immune amplifying effects of cytokine interleukin-1. Dysregulation of the cytokines related to the IL-1 family can lead to pathological derangements because of their influence on inflammation and immunity. One of the components of the immunoglobulin group is the IL-1 receptor (IL-1R). The IL-1R consists of two subtypes namely: subtype 1 (IL-1RI) and subtype 2 (IL-1RII). Subtype 1 (IL-1RI) is present on T cells, endothelial cells, keratinocytes, hepatocytes, and fibroblasts which binds IL-1α preferentially. Subtype 2 (IL-1RII) is a preferred ligand for IL-1β and is generated in bone marrow cells, B cells, neutrophils, and monocytes. Macrophages and monocytes produce interleukin-1 beta (IL-1β) and interleukin-1 alpha (IL-1α) during defensive reactions that occur in the human body when inflammation is detected. Eleven structurally similar proteins that constitute components of the IL-1 superfamily are all involved in inflammation for its regulation [16]. Acute phase of inflammation is caused by IL-1, which results in both systemic and local reactions, including fever, vasodilation, hypotension, and pain sensitivity. Furthermore, it stimulates endothelial cells to express adhesion molecules, which permits immune components and inflammatory cells to penetrate the tissues. Proinflammatory cytokines stimulate keratinocytes in the epidermis to produce interleukin-1(IL-1), which is primarily secreted by monocytes, tissue macrophages, and dendritic cells. B lymphocytes, natural killer (NK) cells, and epithelial cells also express IL-1, with the stratum corneum being the major source of active IL-1 [17]. IL-1 molecules are not just associated with inflammation, despite the fact that they are typically thought of as basic proinflammatory cytokines. Additionally, they have been demonstrated to be essential to a wide range of other physiological and pathological processes, such as cognitive regulation, sleep, depression, pregnancy, blood cell production, metabolism, and many others [18]. Single nucleotide polymorphism (SNP) is a single genomic variant that is found in the DNA between genes which plays a crucial role in the field of medicine as these are associated with disease and acts as biological markers. The researchers aim to focus on the specific genetic variants known as IL1RA_mspa11,100_(rs315952) in finding the association of the alleles to the systemic lupus erythematosus.

Meanwhile, the IL1RA_mspa11,100_(rs315952), a single nucleotide polymorphism (SNP) in the 1L-1RN gene that encodes interleukin-1 receptor antagonist (IL-1Ra), shows an association with several health conditions revolving around inflammation and immune responses [19]. Their findings noted that the SNP itself is linked to a lower risk of having acute respiratory distress syndrome (ARDS) occurrence. This polymorphism is also associated with the increase in IL-1Ra plasma levels, which indicates protective roles in inflammatory conditions. The polymorphism also relates to obesity-related hypertension, which may pose a potential impact on metabolic and inflammatory pathways [20].

It has also been suggested that the IL1RA_mspa11,100_(rs315952), a cytokine gene polymorphism of the IL-1R, is also implicated in autoimmune diseases. It was implied that variations in the mentioned gene, which was influenced by the rs315952 polymorphism, can affect the immune response in SLE [21]. Additionally, Iranians showed that the T allele of the polymorphism may affect patients’ risk of autoimmune conditions, as the polymorphism alters the IL-1Ra activity results [22]. Interestingly, it was noted that the concept of the rs315952 polymorphism to an increased risk of SLE is distantly related [23]. These had paved the way for the inconsistencies of their findings. In some SNPs of the IL-1R like the IL1R1_pstl 1970_(rs2234650), this SNP had not been correlated to an increased risk of SLE [<u>22</u>; <u>23</u>; <u>24</u>].

Recent studies explored the various mechanisms that associate the biomarkers of the IL-1R receptor with risk of SLE occurrence. Among them asserts that the IL-1R family of soluble receptors are biomarkers suitable for indicating the presence of SLE [25]. Moreover, the enhanced expression of the IL-1R can indicate inflammatory symptoms for SLE [26]. Under the IL-1R gene polymorphisms enlist possible combinations of varying alleles which include the genotypes C/C, C/T, and T/T. Among them, the T allele is usually found among SLE patients, which may potentially indicate that the T allele situated within the IL-1R gene corresponds to an increased risk of SLE [22]. In the context of the IL-1RN gene, there were varying alleles within the gene polymorphisms of rs315952, at which patients carrying the T allele were associated with various hematological manifestations including neutropenia. The clinical presentations of neutropenia are profound. In general, neutropenia in SLE is linked to an increased risk of various infections, notably during disease flares when neutrophil counts exhibit the possibility of a significant drop [27]. This case is common in patients with higher disease activity, based on the Systemic Lupus Erythematosus Disease Activity Index (SLEDAI) [28]. Due to these risks, infections lead the cause for morbidity and mortality of SLE patients [<u>27</u>; <u>29</u>]. Therefore, neutropenia can be utilized as a marker for disease activity as prompted by IL-1RN gene polymorphisms. Lower neutrophil counts may indicate more severe manifestations of SLE [<u>27</u>; <u>30</u>]. All these imply that the cytokine gene polymorphisms of the IL-1RN gene involving the rs315952 at locus Mspa-11,100 might suggest its role in the pathophysiology of SLE [24].

In light of these, studies suggested that cytokine gene polymorphisms (as a whole), suggest its ties with the incidence of SLE due to its capacity to trigger inflammation, and immune disruption [<u>31;32</u>]. However, the varying genotypes of these cytokine gene polymorphisms can still be left uncertain, and may leave room for further research. Interestingly, few studies still suggest the association of the selected IL-1RN cytokine gene polymorphisms with the risk of SLE [23]. This implies that due to the insufficiency of data, little is still known as to whether or not the cytokine gene polymorphisms of rs315952 is a risk factor for SLE due to the inconsistencies with the findings and limited studies.

Majority of the studies had been clustered around centering on providing the implications that these cytokine gene polymorphisms had for the bodily system, and not entirely on the concept of whether or not these cytokine gene families establish links with certain autoimmune diseases, especially with SLE. Considering these, the relationship and correlation between the IL-1RA cytokine gene polymorphism and SLE is distantly grasped. As a result, this meta-analysis looks into the association of the IL-1RA cytokine gene polymorphism with SLE, and as well as the alleles that comprise these. The primary purpose of this meta-analysis paper seeks to answer and give light with regards to the uncertainties of the results pertaining to the associations of IL-1RA cytokine gene polymorphism with SLE occurrence. By synthesizing the findings of all accessible papers, this meta-analysis sought to yield more accurate and precise estimates in order to ensure the robustness of the paper, while considering the statistical power held by each study.

## II. RESEARCH OBJECTIVES

1. Assess the gene variants (polymorphisms) of the IL-1RA cytokine gene (rs315952) and foresee possible patterns towards developing a risk for systemic lupus erythematosus (SLE) among Asians by utilizing the following genotypes:

- C/C
- C/T
- T/T
2. Assess the individual alleles of the IL-1RA cytokine gene (rs315952) and foresee possible patterns towards developing a risk for systemic lupus erythematosus (SLE) among Asians by utilizing the following alleles:

- C
- T

## III. METHODS

### Search Strategy

The combination of English key terms utilized for searching were “Il-1R polymorphisms Il-1R, interleukin-1 receptor and SLE, rs315952 and SLE, Il-1R cytokine gene polymorphisms and SLE, IL-1RA_mspa11,100_ (rs315952) and Systemic Lupus Erythematosus, Il-1R gene polymorphisms and SLE, IL-1RA_mspa11,100_ (rs315952) and SLE, Il-1R gene polymorphisms and systemic lupus erythematosus, Il-1R cytokine gene polymorphisms and systemic lupus erythematosus, IL-1RA_mspa11,100_ and SLE *”.* Literature search and article selection were carried out in PubMed, Springer, Cochrane Library, and as well as search engines like EBSCOHost Platform and Google Scholar were extensively searched for data mining. The respective titles and abstracts were initially screened; afterwards, a thorough evaluation of the full paper followed. The significant references that were cited from the successfully screened papers were also screened to identify potential studies to be included.

### Inclusion and Exclusion Criteria

If the specified criteria were met, researchers regarded the studies considered to be eligible for inclusion: (1) A case control and cross-sectional study featuring the patients manifesting SLE (cases), and those that were not (healthy controls); (2) The SLE diagnosis of case patients were based on the revised criteria of the American College of Rheumatology; (3) A polymerase chain reaction (PCR) and a polymerase chain reaction-specific sequence primer (PCR-SSP) was utilized for cytokine gene typing; (4) The PCR products of the IL-1R cytokine gene were subjected to electrophoresis on a 2% agarose gel; (5) A Hardy-Weinberg equilibrium was utilized for a comparison between the observed and expected genotypic frequencies among the SLE patients, and the healthy controls; (6) Papers showing the allele frequencies for IL-1RA_mspa11,100_ (rs315952) on case and control patients; (7) Paper comprising of a sample size of at least 75 subjects; and (8) Articles that were written and published in the English language. Moreover, citations from the studies that were chosen were additionally looked through for other suitable articles.

An exclusion criteria was also formulated, and may result in a screened study as negligible for data mining. Thereby, an automatic exclusion of the paper ensues once it contains any of the following criteria: (1) Only one SNP of the target variant was enlisted, and no other included genotypes summarized in a table were visible per se.; (2) Longitudinal and cohort studies; (3) Papers enlisting an Odds ratio with a confidence interval of less than 95%; (4) Studies that do not offer the entire content, permitting insufficient information regarding their methodology, statistical analysis, and research design.

### Data Extraction

For each eligible study that was successfully screened, the following data was taken out: the last name of the first author, the year of publication, the participants’ country and ethnicity, the total number of participants, the number of case (SLE patients) and control (healthy patients), and the corresponding genotypes (C/C, C/T, T/T) of IL-1RA_mspa11,100_(rs315952).

### Methodological Quality assessment of the included studies

The Newcastle-Ottawa Scale (NOS) was utilized for the quality assessment of the eligible studies. The selected studies had undergone careful assessment using the following parameters: selection, comparability, and outcome. All studies utilized for the study were graded as fair-good quality under the AHRQ standards. All successfully screened studies that garnered 5-9 stars at NOS were included.

### Statistical analysis

The statistical analysis for this meta-analysis paper utilized a Review Manager 5.4.1. which is a software analysis used for the construction of forest plots in order to assess possible overlapping confidence intervals that are present within the eligible studies. In addition, the odds ratio that were pooled, and as well as the 95% study and total confidence intervals were obtained using the fixed and random effects models in order to explore the degree of heterogeneity among varying studies. Specifically, the I2 statistics was carefully checked for the detection of heterogeneity as 0-40% deems the degree of heterogeneity as negligible. Any forest plot constructed with an I2 statistics of greater than 40% will prompt for a recheck of the data that was gathered, and also consider the exclusion of outlier studies. After the construction of the final forest plot pooled odds ratio concludes, the researchers analyzed whether the alleles and genotypes exhibit significant associations with SLE.

### Sensitivity analysis and publication bias

Publication bias and outlier determination was assessed through a funnel plot asymmetry offered by the Review Manager 5.4.1. software. Regardless, all studies utilized will be graded as fair-good quality under the AHRQ standards. All successfully screened studies that garnered 5-9 stars at NOS will be included. Aside from these, Sensitivity analysis is an analysis used to determine the robustness of the study. For this meta-analysis, omitting one study at a time will be incorporated in order to measure the robustness of the study.

## IV. RESULTS

### Search Results

Figure 1 summarizes the study selection process utilizing Preferred Reporting Items for Systematic Reviews and Meta-Analysis (PRISMA) guidelines [33]. The initial search yielded a total of sixty-six (66) studies that were thoroughly assessed. The titles of the paper and articles that clearly stated the possible relationships between the IL1RA cytokine gene and systemic lupus erythematosus will be evaluated by the checklist stated at figure 1. Following the set of criteria, after removing redundant and unnecessary research, six (6) publications in all satisfied the inclusion criteria for this systematic review and meta-analysis (Tsai et al., <u>21</u>; Tsai & Lan., <u>34</u>; Tahmasebi et al., <u>22</u>; Ziaee et al., <u>24</u>; Dar et al., <u>35</u>; Ahmed et al., <u>23</u>). Other studies that were not included for the study were also documented, and be provided along with the reason for not utilizing it. A study selection process was also presented based on the PRISMA 2020 statement. If a difference in opinion persists, then the article shall be sent to a group of experts using the same checklist for their decision. In addition, a Newcastle-Ottawa Scale (NOS) was also utilized for quality checking of all the screened papers and articles.

**Fig. 1.**
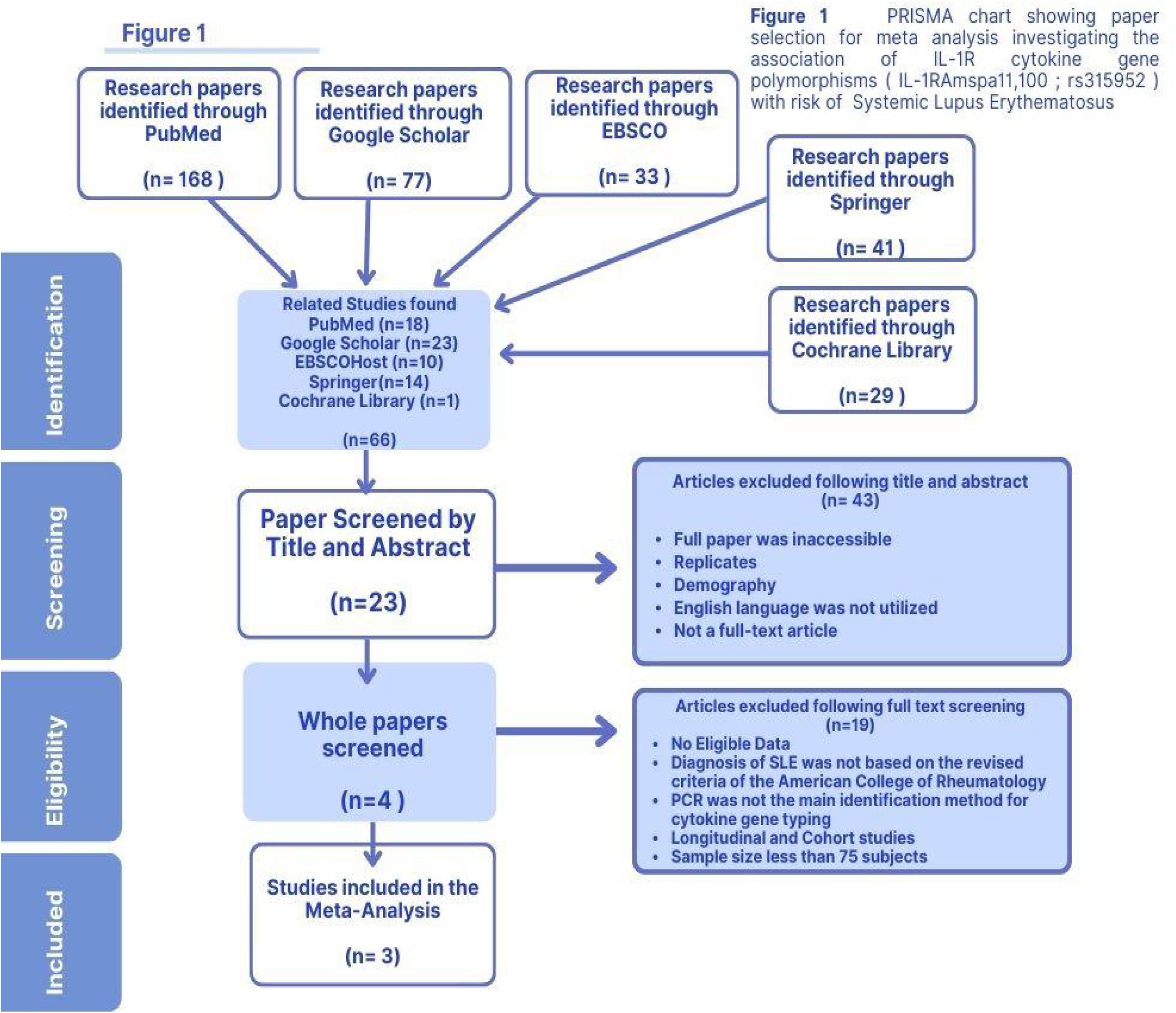
Literature search summary following the PRISMA guideline.

On the contrary, figure 2 shows the characteristics of the included studies. Ideally, most of the publication dates were set 10 years before the year 2024. However, since there is paucity of data, and that journals usually require payment to access their manuscript, the year of publications utilized for this paper was adjusted and ranged from 2006, followed by 2009, and up until 2020. All eligible studies contained case-control groups that explored a specific population in order to determine the association between selected Il-1R cytokine gene polymorphisms and SLE. In addition, the sample size contained as much as 1,309 subjects from all the eligible studies tallied.

**Figure 2.**
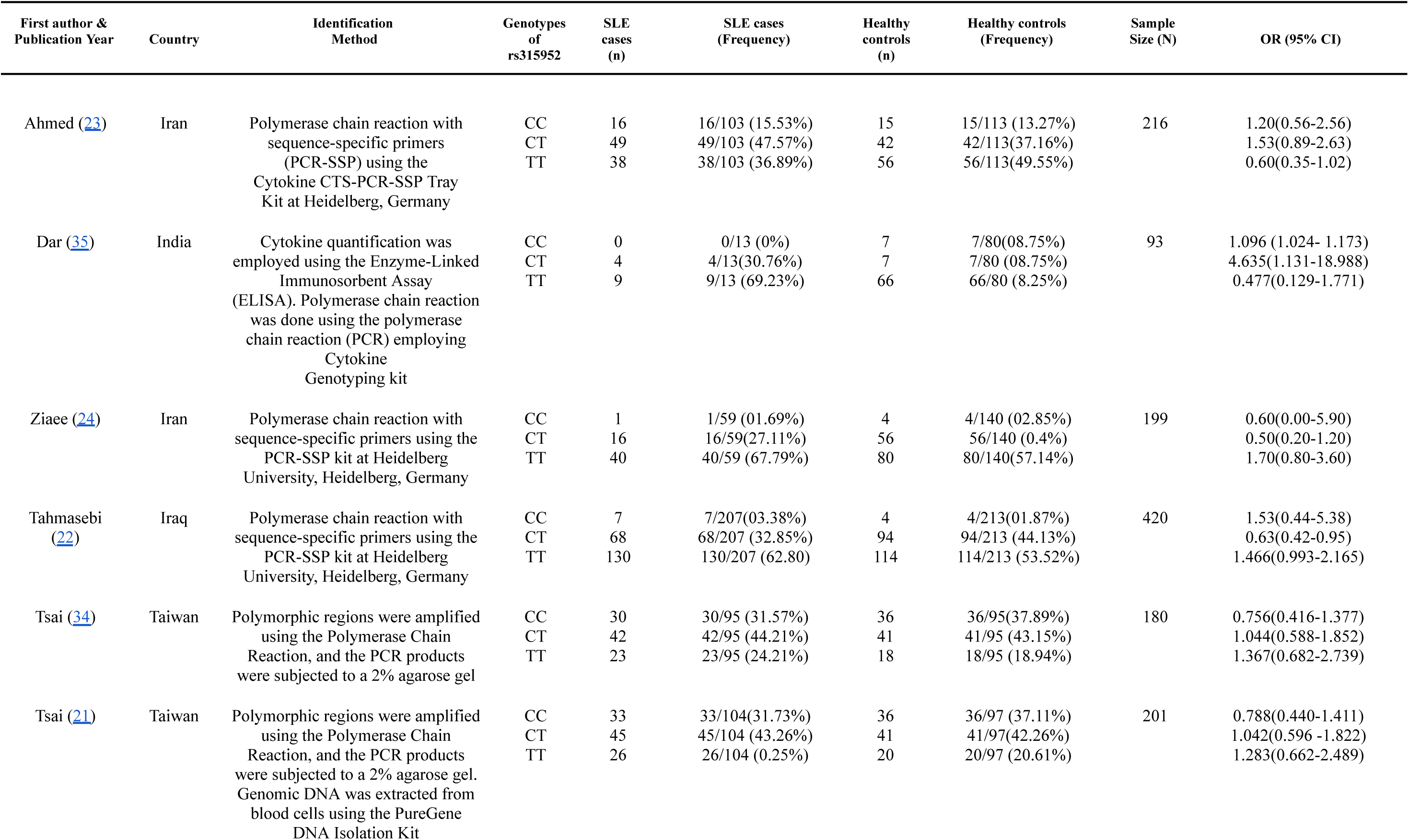
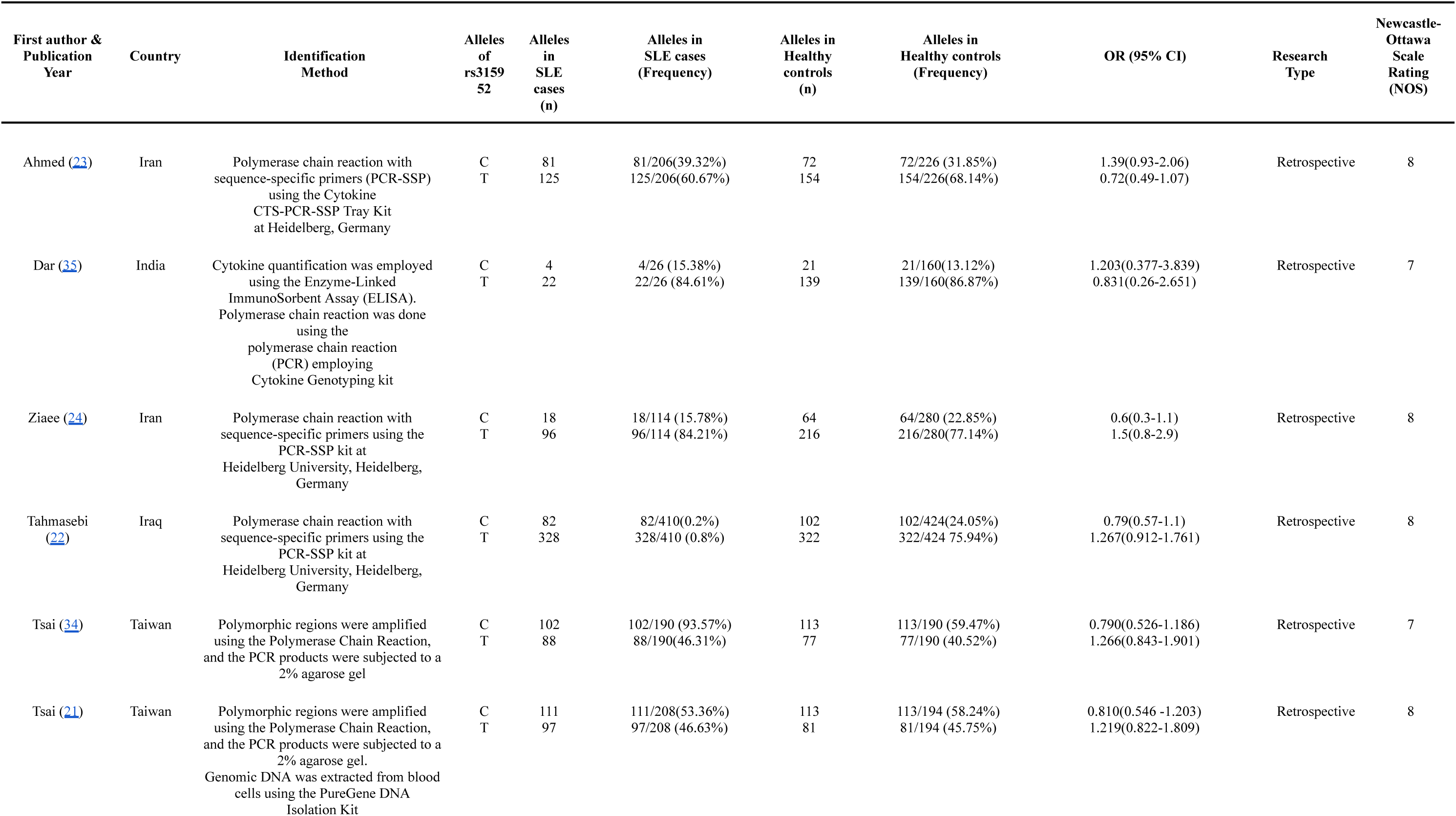
Genotypes of rs31592 among SLE Patients and Healthy Controls

### Overall analysis for the association of the Il-1RA cytokine gene polymorphism and SLE

The six (6) included studies provided the list of genotypes for IL-1RA_mspa11,100_ (rs315952), as well as the summation of all the C and T alleles. The overall pooled analysis suggests that the **T/T genotype**, as well as the individual **T allele** have **significant associations** with the risk of SLE. On the other hand, the C/C, and the C/T genotype, and the C allele show no significant association with the risk of the disease. This implies that there is an increased likelihood that a patient may develop SLE whenever the genotype T/T and the T allele exist.

The fixed effects model (Fig 3.1) displayed **no significant associations** on the C/C genotype towards SLE risk as due with the pooled OR being less than 1 (OR 0.88; CI 0.63-1.24; P^A^= 0.48). In accordance with these values, this also concludes that the effects were **statistically significant**. The figure’s pooled analysis showed homogeneity (I2 = 0%, P^H^ = 0.73), which indicates the combinability of the individual studies. As per the pooled ORs of the selected studies, individuals with the C/C genotype for IL-1RA_mspa11,100_ (rs315952) are 12% less likely to have SLE. In line with this, a funnel plot (Figure 3.2) for Figure 3.1 was formulated, which revealed no outlier among the studies, indicating the combinability of the individual studies as homogeneity was presented, indicating high consistencies that the result gathered was certain.

**Figure 3.1.**
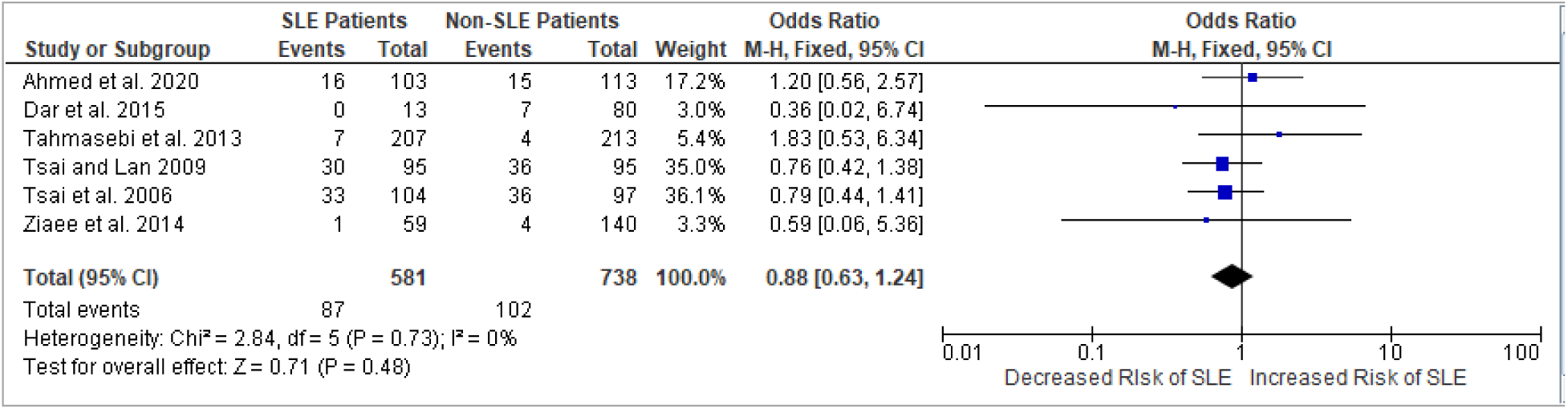
Forest plot association between the C/C genotype in rs315952 of SLE and non-SLE patients

**Figure 3.2.**
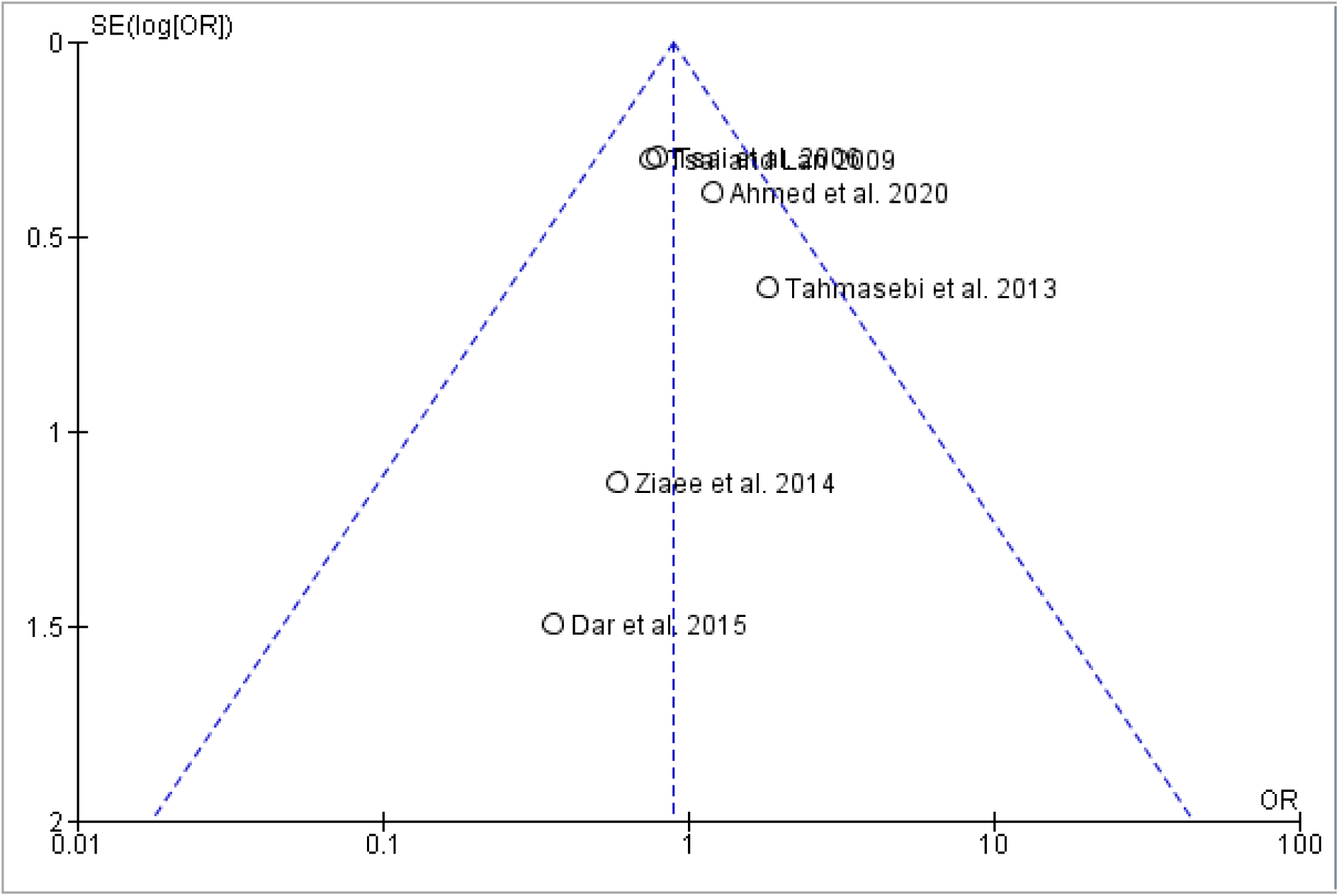
Funnel plot of Figure 3.1 for determination of possible outlier studies

A funnel plot (Figure 4.2) based on the analysis of the association between the C/T genotype of rs315952 and SLE (Figure 4.1) was formulated to identify outliers. The said plot exhibited the studies of [22] and [35] as outliers, which led to their exclusion in the final analysis. As a result, the final fixed effects model (Fig 4.3) displayed **no significant** association between the C/T genotype and the risk of SLE (OR 0.76; CI 0.59-0.98; P^A^= 0.03), declaring that the results were **statistically significant**. The figure’s pooled analysis showed moderate heterogeneity (I2 = 29%, P^H^ = 0.24), which indicates the combinability of the individual studies with a slightly strong conclusion. Additionally, the pooled ORs of the selected studies indicate that individuals with the CT genotype for IL-1RA_mspa11,100_ (rs315952) are 24% less likely to have SLE.

**Figure 4.1.**
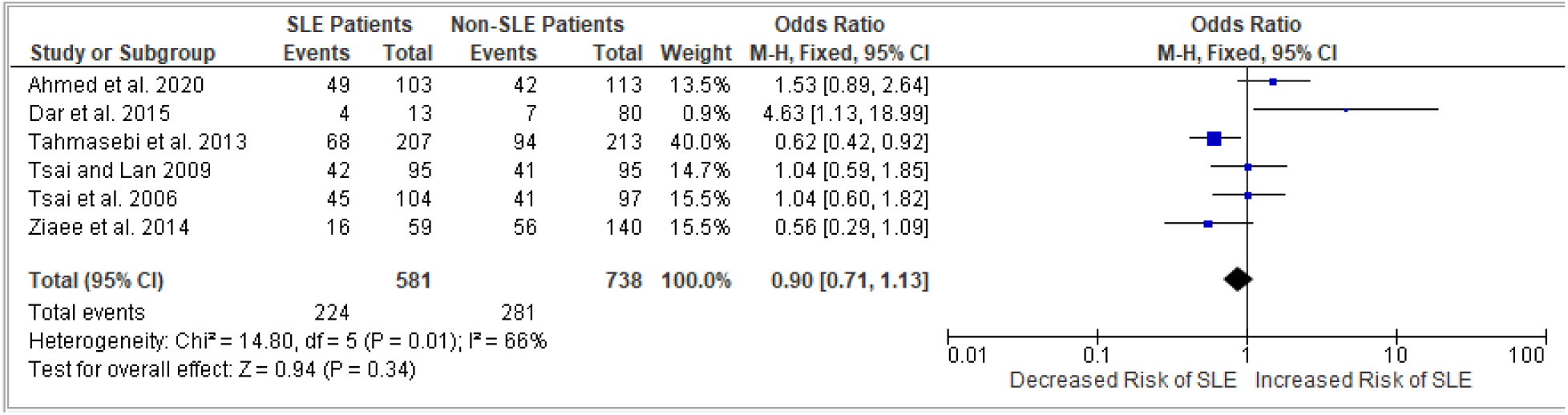
**Initial** forest plot association between the C/T genotype in rs315952 of SLE and non-SLE patients

**Figure 4.2.**
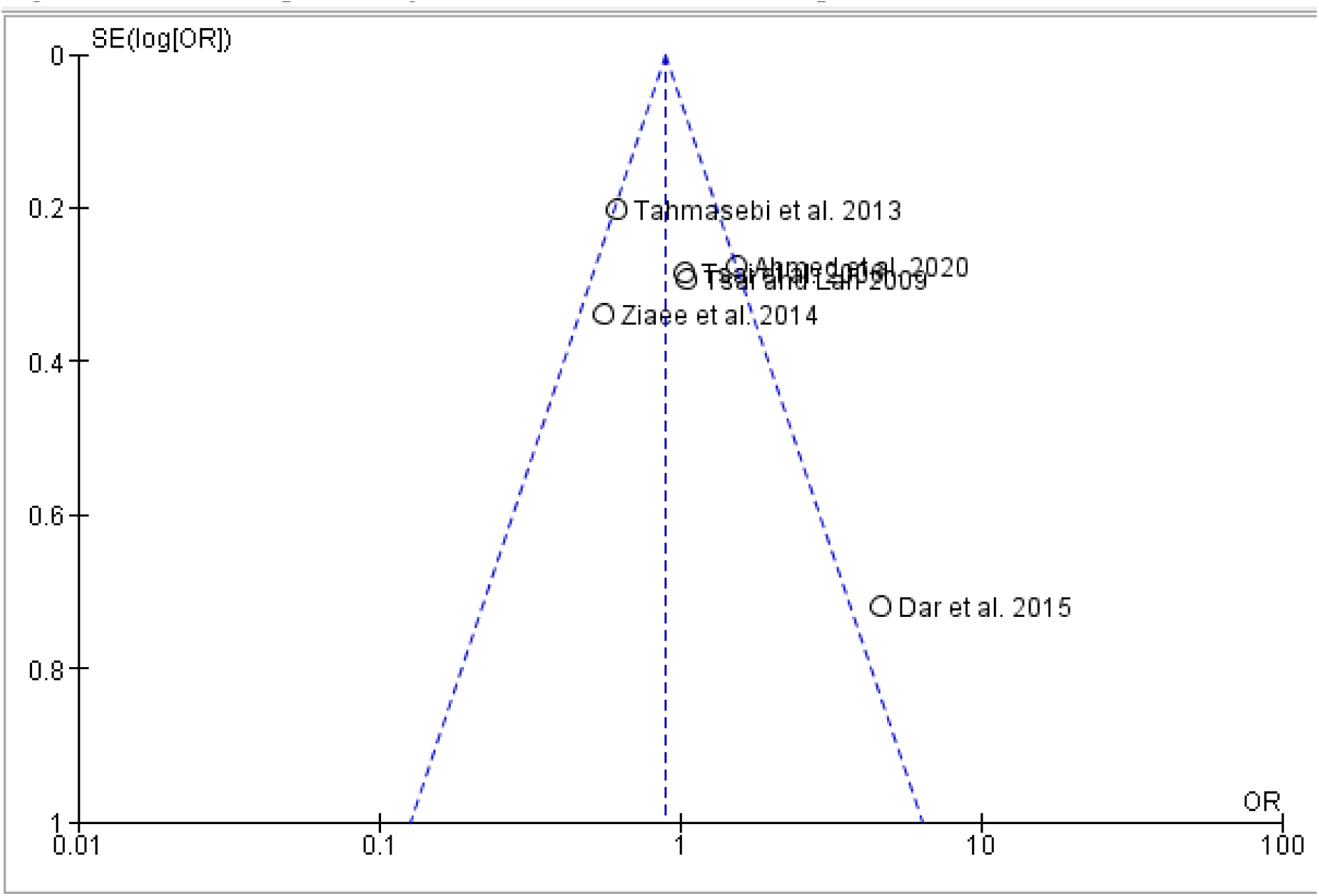
Funnel plot of Figure 4.1 for determination of possible outlier studies

**Figure 4.3.**
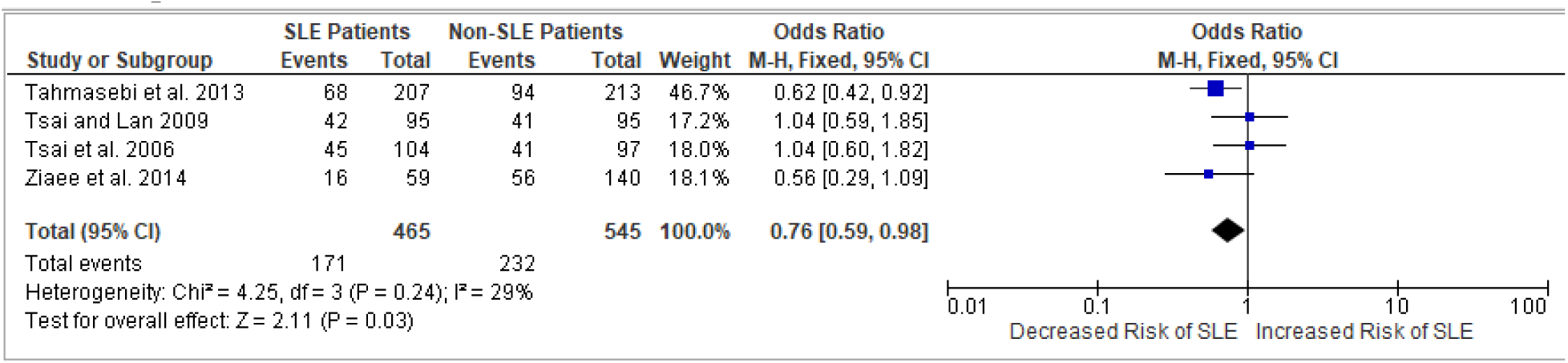
**Final** forest plot association between the C/T genotype in rs315952 of SLE and non-SLE patients

Meanwhile, a funnel plot (Figure 5.2) was also formulated based on the analysis of the association between the T/T genotype of rs315952 and SLE (Figure 5.1) for outlier identification. The said plot pinpointed the study of [23] as an outlier, which led to its exclusion in the final analysis. The final fixed effects model (Fig 5.3) displayed **significant** associations between the T/T genotype and the risk of SLE (OR 1.38; CI 1.05-1.80; P^A^= 0.02). With the pooled ORs being more than 1 (OR>1), this alone implies that **no significant differences** were evident with the results. The figure’s pooled analysis showed homogeneity (I2 = 0%, P^H^ = 0.02), which indicates the combinability of the individual studies.

**Figure 5.1.**
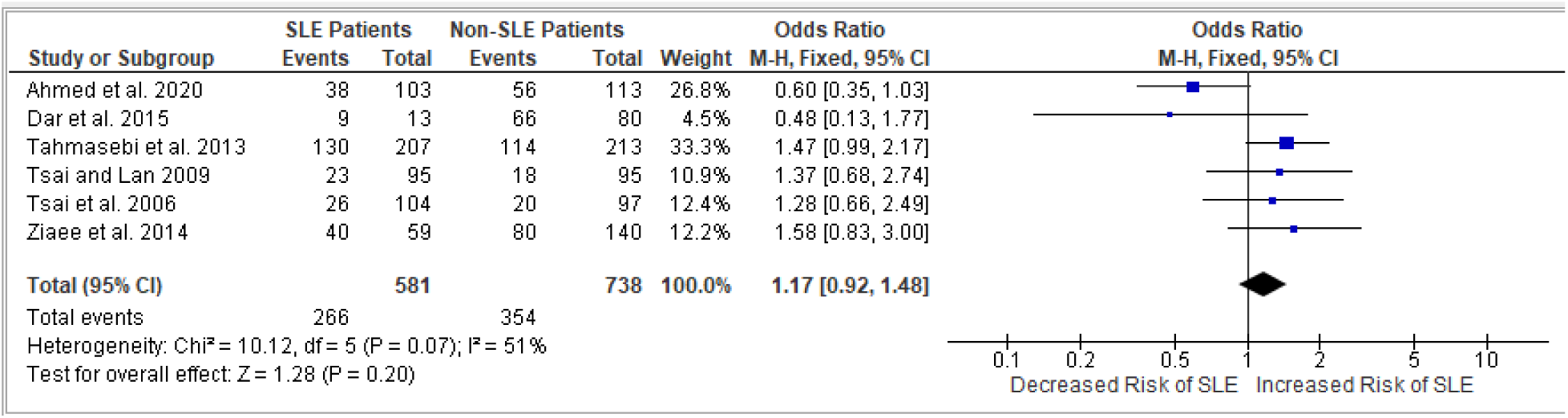
**Initial** forest plot association between the TT genotype in rs315952 of SLE and non-SLE patients

**Figure 5.2.**
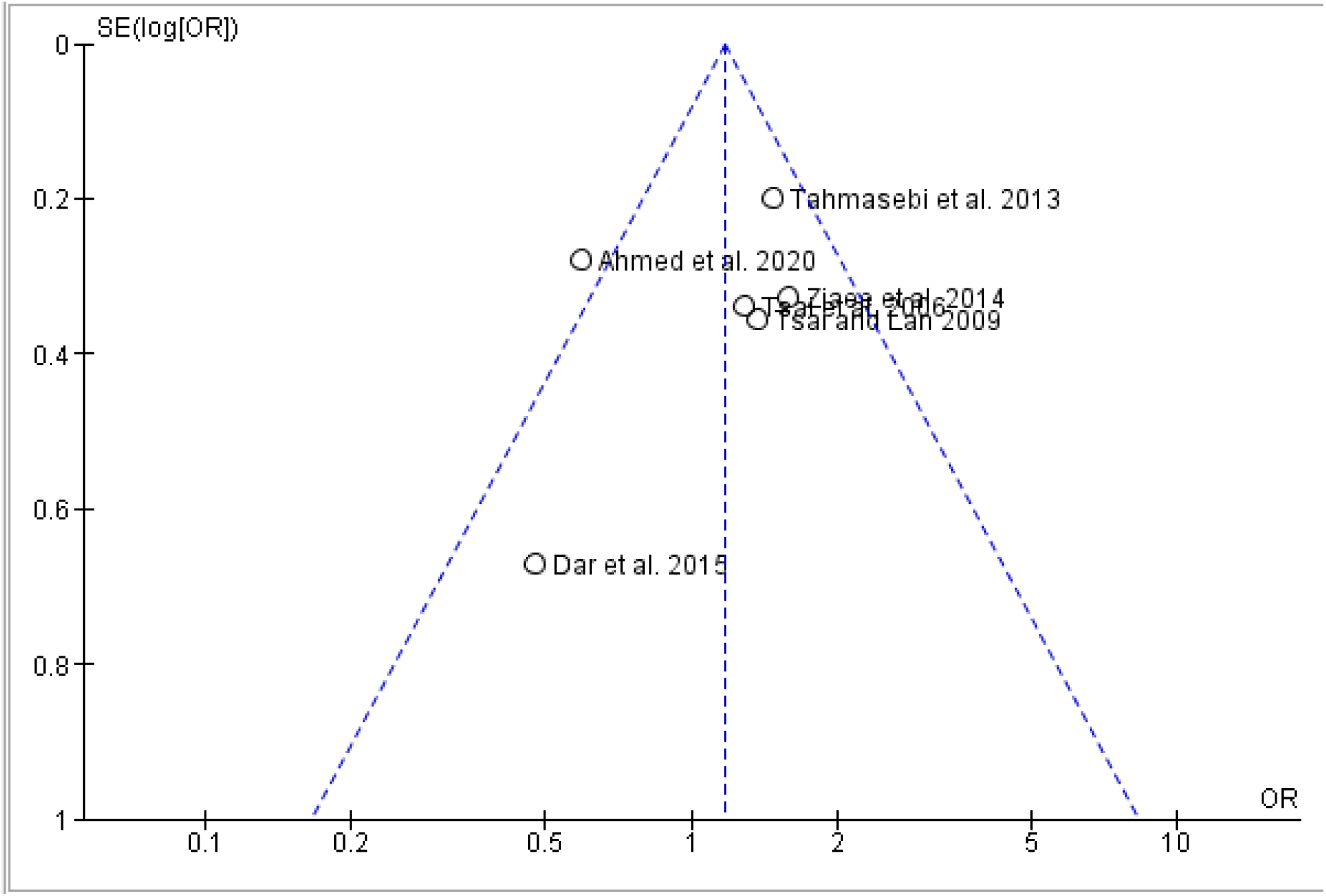
Funnel plot of Figure 5.1 for determination of possible outlier studies

**Figure 5.3.**
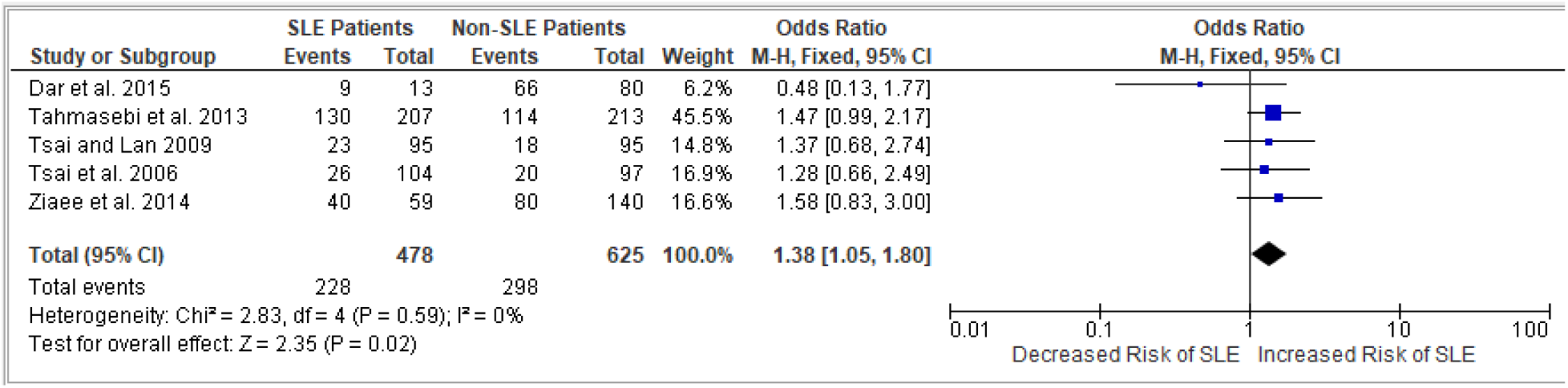
**Final** forest plot association between the TT genotype in rs315952 of SLE and non-SLE patients

The researchers also formulated a funnel plot (Figure 6.2) based on the analysis of the association between the C allele of rs315952 and SLE (Figure 6.1) to identify outliers. The said plot exhibited the study of [23] as an outlier, which led to its exclusion in the final analysis. The final fixed effects model (Fig 6.3) displayed **no significant** associations between the C allele and the risk of SLE (OR 0.78; CI 0.64-0.95; P^A^= 0.01), marking these results as **statistically significant**. The figure’s pooled analysis showed homogeneity (I2 = 0%, P^H^ = 0.90), which indicates the combinability of the individual studies. As per the pooled ORs of the selected studies, individuals with the C allele for IL-1RA_mspa11,100_ (rs315952) are 22% less likely to have SLE.

**Figure 6.1.**
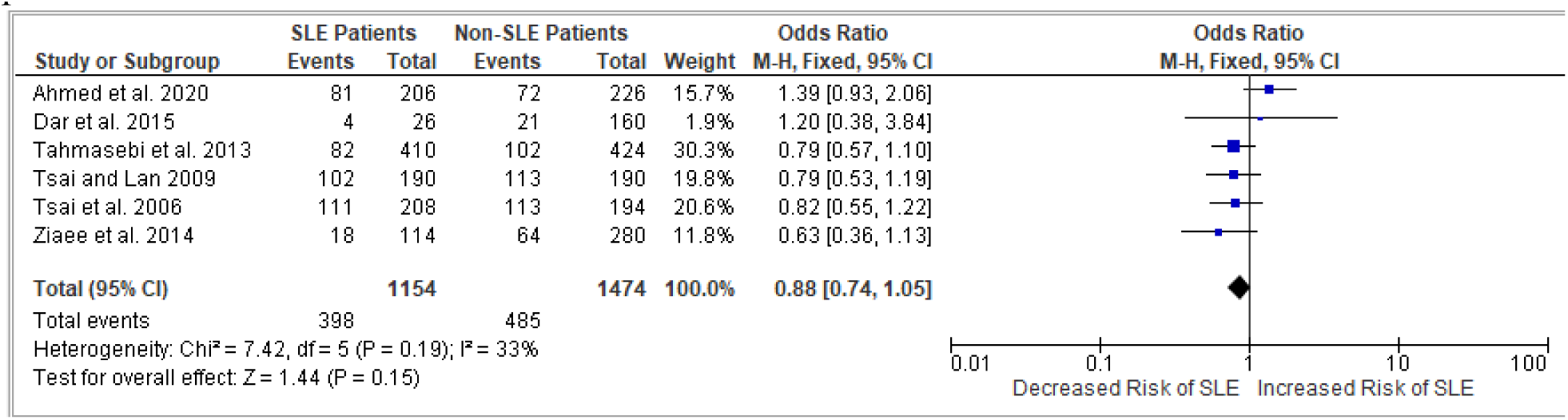
**Initial** forest plot association between the C allele in rs315952 of SLE and non-SLE patients

**Figure 6.2.**
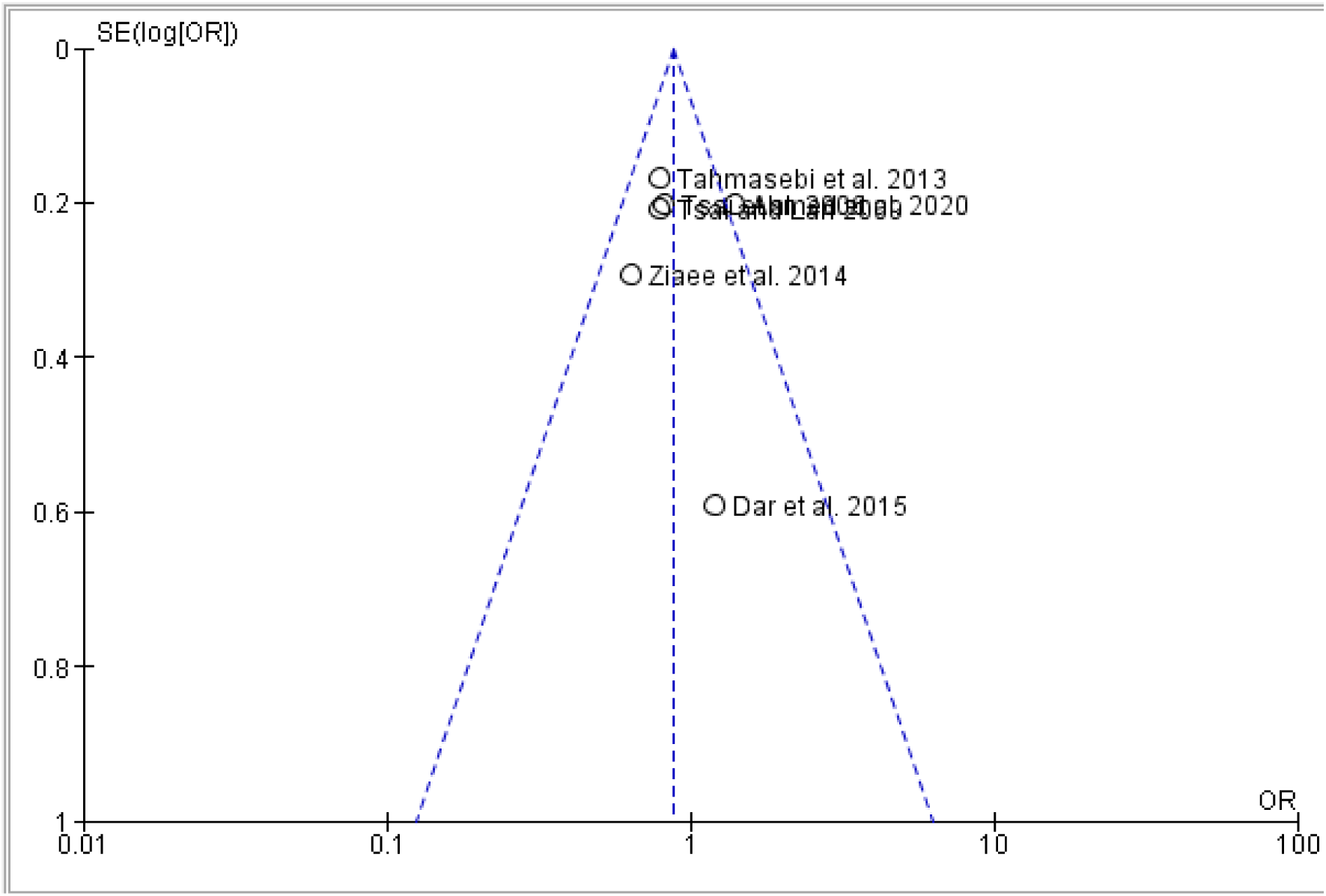
Funnel plot of Figure 6.1 for determination of possible outlier studies

**Figure 6.3.**
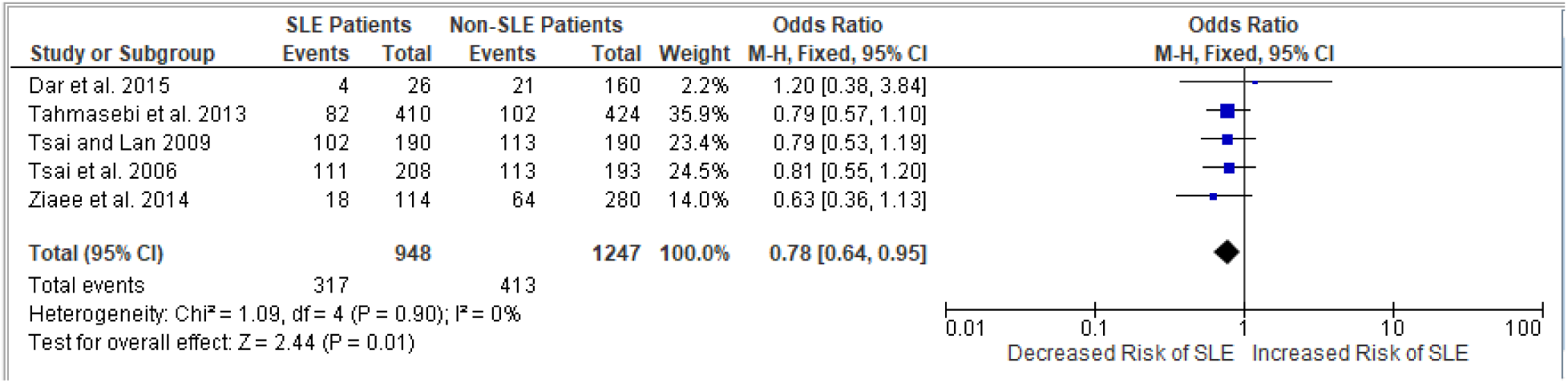
**Final** forest plot association between the C allele in rs315952 of SLE and non-SLE patients

Lastly, another funnel plot (Figure 7.2) based on the analysis of the association between the T allele of rs315952 and SLE (Figure 7.1) was formulated to identify outliers. The said plot exhibited the study of [23] as an outlier, which led to its exclusion in the final analysis. The final fixed effects model (Fig 7.3) displayed **significant association** between the T allele and the risk of SLE (OR 1.28; CI 1.05-1.55; P^A^= 0.02). This also suggests that the results were **not statistically significant.** The figure’s pooled analysis showed homogeneity (I2 = 0%, P^H^ = 0.89), which indicates the combinability of the individual studies.

**Figure 7.1.**
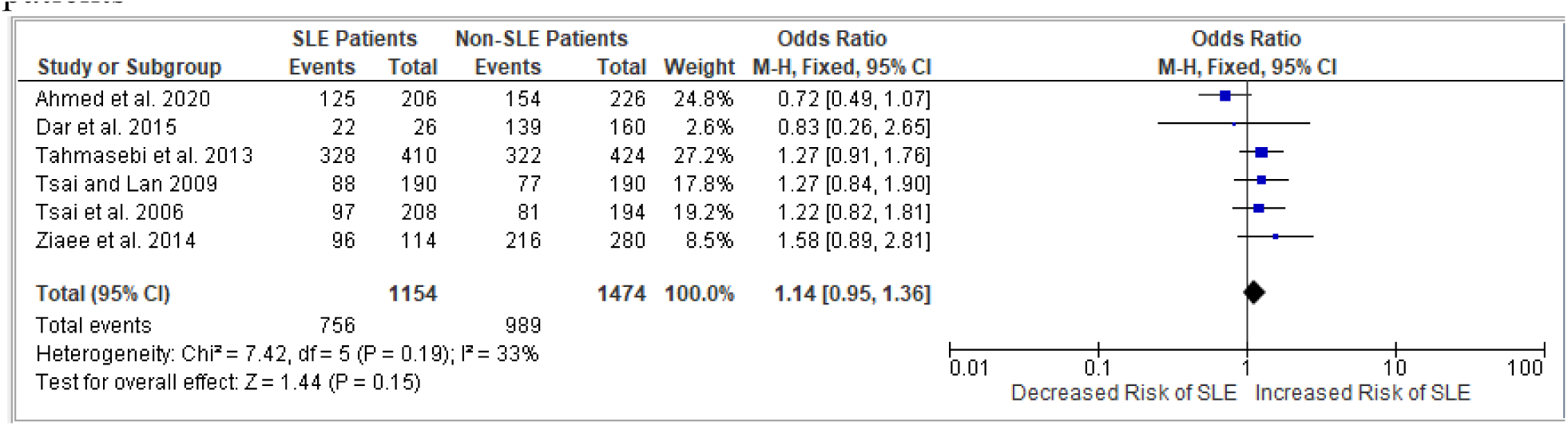
**Initial** forest plot association between the T allele in rs315952 of SLE and non-SLE patients

**Figure 7.2.**
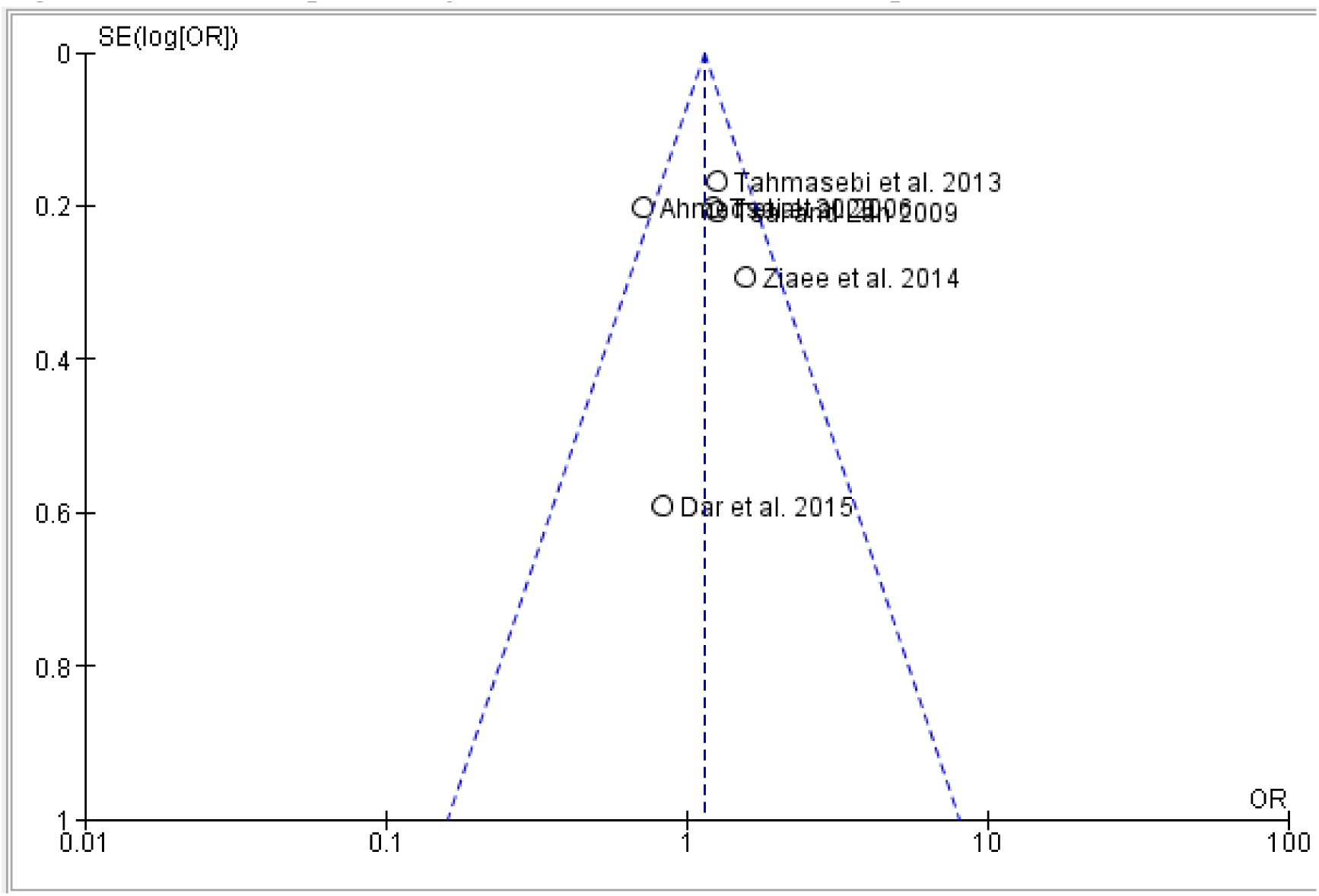
Funnel plot of Figure 7.1 for determination of possible outlier studies

**Figure 7.3.**
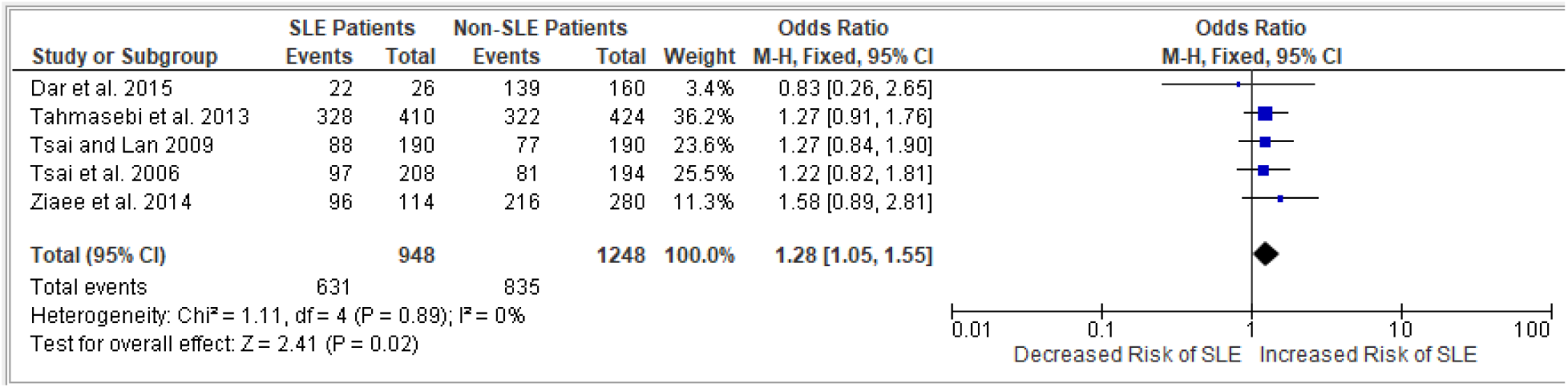
Final forest plot association between the T allele in rs315952 of SLE and non-SLE patients

## V. DISCUSSION

IL-1R cytokine family of proteins are proteins that are involved in cell signaling, and a dysregulation of some of these proteins may permit certain polymorphisms being candidates for an increased risk of certain autoimmune diseases, such that of SLE (<u>19</u>; <u>21</u>). The current meta-analysis paper explored IL1RA_mspa11,100_(rs315952) as to whether or not this SNP induces an increased risk of SLE.

The findings to the present meta-analysis study provided strong evidence of the potential association between the rs315952, and SLE emergence. This evidence had taken into account the robustness of the study by omitting studies that were deemed as outliers for a specific allele and genotype. In line with these, homogeneity of the results were also assured in order to draw accurate conclusions (I^2^ = 0-29%). Furthermore, this meta-analysis’s quality of evidence is strengthened by a high degree of significance, consistent effect precision, and reliable post-outlier outcomes. With the present study synthesizing all the results from 6 studies, a combined total of 1,309 participants were gathered.

Among the findings, the C allele (OR: 0.78; 95% CI: 0.64–0.95; P^A^: 0.01; I^2^ = 0%), and as well as the C/C (OR: 0.88; 95% CI: 0.63–1.24; P^A^: 0.48; I^2^ = 0%) and the C/T (OR: 0.76; 95% CI: 0.59–0.98; P^A^: 0.03; I^2^ = 29%) genotypes of the rs315952 indicated that there was a less likelihood of developing SLE whenever these genotypes were acquired among SLE and non-SLE individuals. However, gaps were evident on the genetic models C/C, C/T, and the C allele of the rs315952 as the effects were statistically significant. These had deviated from the normal findings as the IL-1R, when defective, were known to induce autoimmune diseases like SLE, regardless of the alleles and genotypes. Thus, the pathogenesis of SLE may remain elusive, and requires further studies. Furthermore, based on the pooled ORs, the current study suggested that the C/T genotype exerted a more protective effect against SLE compared with the C/C genotype since the study had found that individuals having the C/T genotype of the rs315952 were 24% less likely to develop SLE, compared with the 12% less likelihood of developing SLE if an individual had a C/C genotype. However, the latter point may not be thoroughly proven as outlier determination for the pooled analysis of the C/T genotype was incorporated.

Moreover, the T allele (OR: 1.28; 95% CI: 1.05–1.55; P^A^: 0.02; I^2^ = 0%) and the T/T genotype (OR: 1.38; 95% CI: 1.05–1.80; P^A^: 0.02; I^2^ = 0%) alone of the IL-1RA_mspa11,100_ (rs315952) can be considered as risk factors in the development of SLE. In some SNPs of the IL-1R such as the IL-1R1_pstl1970_(rs2234650), even if the T allele and the T/T genotype was acquired, a negative association with SLE was traced, implying that there were genetic variations that permit an increased disease susceptibility to the onset of SLE (<u>22</u>; <u>23</u>; <u>24</u>). Nonetheless, the T allele and the TT genotype were consistent with what had been known for SLE emergence.

### Comparison with other studies

The current meta-analysis’ findings were consistent with those of the previous studies (Tsai et al., <u>21</u>; Tsai & Lan., <u>34</u>; Tahmasebi et al., <u>22</u>; Ziaee et al., <u>24</u>; Dar et al., <u>35</u>; Ahmed et al., <u>23</u>). In accordance with [23], the IL-1RA_mspa11,100_ (rs315952) genotypes, including the C (OR: 1.20) and T (OR: 0.72) alleles, and as well as the C/C (OR: 1.20), C/T (OR: 1.53), and T/T (OR: 0.60) all showed opposing outcomes in this study. This had led to the study being omitted for most cases in order to ensure minimal heterogeneity. There were also slight deviations garnered from other studies compared with the study of [35], at which the study suggested an increased risk of SLE for the C/T genotype (OR: 4.63; 95% CI: 1.13-18.99). Other deviations from the findings were also evident; however, these studies were not omitted for a certain figure as bases from the I^2^ statistics, and as well as the basis of the funnel plot for outlier determination.

### Limitations and potential biases of the study

The overall findings of the current paper suggest the association of the polymorphisms of the IL1RA_mspa11,100_(rs315952) SNP and its possible correlations to an increased susceptibility of SLE. However, it should be noted that limitations arise while interpreting the findings which included (1) the diversity of the year of publications starting from the year 2006 up until 2020 which indicates that some studies may be deemed to be outdated if the 10-year mark of citing sources was abided; however, due to the limited data, and with some journals requiring paid access, the study years below 2014 was deemed relevant (for as long as the publication year will not be published during the 1990s). Aside from that, the study was not able to provide a (2) representation of the gender distribution between SLE and healthy controls (as SLE is more renowned for women, and perhaps, the trend may be different compared with men). Additionally, (3) different ethnic groups from the different countries were not emphasized. (4) Failure to include additional systemic lupus erythematosus risk factors and other genotypes. Considering these limitations, the findings of this meta-analysis need to be evaluated cautiously when utilized in a medicinal setting. This meta-analysis, to the extent of the researcher’s knowledge, examines the association between the IL-1RA_mspa11,100_ (rs315952) cytokine gene polymorphism and the risk of systemic lupus erythematosus. With this approach, the researchers aim to contribute to the understanding of the pathophysiology of systemic lupus erythematosus by identifying risk factors for this autoimmune disease through means of associating it with the rs315952 cytokine gene polymorphism. This offered an innovative approach to determining and identifying the occurrence of SLE based on its genetic factor. Moreover, based on the resulting pooled ORs, the C allele, and as well a the C/T and C/C genotypes acquired among SLE and non-SLE individuals have less likelihood of developing SLE, while the pooled ORs for the T allele, and the TT genotype suggest that it can be considered as risk factors in the development of SLE.

## VI. CONCLUSION

A general retrospective analysis of the respective genetic models C/C, C/T, and T/T genotypes, as well as the examination of the C and T alleles of the IL-1RA_mspa11,100_ (rs315952) cytokine gene polymorphism suggested varying outcomes to SLE risk. While the T allele and the TT genotype strictly abides with the current findings highly linked with SLE emergence, still, the C/C, C/T, and the C allele showed contrasting findings, which suggested that the results were statistically significant, implying that an individual carrying at least a single copy of the C allele exerts a protective effect against SLE. With regards to the C/T genotype, the exclusion of the studies [22] and [35] prompted for a finding that also strongly suggests a protective effect against SLE. The exact etiology behind all these processes still remains elusive, and rigorous studies are required to determine the type of inheritance that the varying genotypes of the IL-1RA cytokine gene polymorphism represent. Additionally, the haplotypes of the subjects at the IL-1RA gene can undermine the findings of the current study as this concerns a wide range of possible alleles that were inherited/acquired all at once. There is also in dire need of a consideration of the environmental condition that each subject was immersed to as it was established that SLE is both an inherited and acquired autoimmune disease. Moreover, epigenetics may also alter the gene and alleles of the patient which may cause slight inconsistencies with the results. Still, the findings of the current study remain certain that a copy of a T allele and a T/T genotype at the IL-1RA_mspa11,100_ (rs315952) implies for an increased risk for SLE susceptibility, and that a copy of a C allele, and the C/C and C/T genotypes located closely along the gene IL-1RA_mspa11,100_(rs315952) suggests a protective effect against SLE. These findings can serve as a springboard for further analyses pertaining to the knowledge on SLE pathogenesis, and still require rigorous research to be utilized in the field of the health sciences.

## Data Availability

All data produced in the present study are available upon reasonable request to the authors

https://www.medrxiv.org/content/The_Association_of_the_IL-1RA_(rs315952)_Cytokine_Gene_Polymorphism_and_Susceptibility_to_Systemic_Lupus_Erythematosus:_A_Meta-analysis.com

## Author contributions

Study Title and Conceptualization: Carlos, Jan Mikkos L.; Revised by Gabriel, Mary Angelei T. Abstract: Gatbunton, Lark Joshua

Introduction: Carlos, Jan Mikkos L., Gabriel, Mary Angelei T., Gatbunton, Lark Joshua

Study Objectives: Carlos, Jan Mikkos L.; Reviewed by Gatbunton, Lark Joshua & Gabriel, Mary Angelei T.

Figure for PRISMA: Gabriel, Mary Angelei T.; Reviewed by Gatbunton, Lark Joshua

Excel file for the Study Screening Process: Gatbunton, Lark Joshua & Gabriel, Mary Angelei T.

Figure for Study Characteristics: Carlos, Jan Mikkos L.; Revised by Gabriel, Mary Angelei T.

Methodology and RevMan 5.4.1. Partial Results: Carlos, Jan Mikkos L.

Narrative of the Study Results: Gabriel, Mary Angelei T., & Gatbunton, Lark Joshua

General Discussion and Limitations: Carlos, Jan Mikkos L. & Reviewed by Gabriel, Mary Angelei T.

Study Comparison with other Studies: Gabriel, Mary Angelei T., & Gatbunton, Lark Joshua

Conclusion: Carlos, Jan Mikkos L.; Reviewed by Gabriel, Mary Angelei T. & Gatbunton, Lark Joshua

## Statement of the Institutional Review Board

The current study protocol was registered on 25th October, 2024, and approved for eligibility by The International Prospective Register of Systematic Reviews under the National Institute for Health and Care Research (ID: CRD42024601017).

## Data Availability

The raw data for the data mining process, and the files for the statistical analyses that support the current study were not publicly made available. However, the authors can be reached out through their emails if in need of a copy concerning the Microsoft Excel file for the screening process, and the RevMan 5.4.1. file enlisting the partial results, provided that the intention for requesting the files were valid.

## Acknowledgements

We would like to thank Sir Melandro G. Cunanan, M.Sc., Ma’am Zenaida F. Mergal M.Sc., and Ma’am Lani G. Tabangay M.Sc. for their unwavering support and guidance throughout the writing process of this study. Their expertise and guidance had directed us by providing valuable feedback and other possible ideas that enabled us to modify and enhance the quality of our paper accordingly. We would also like to thank Sir Raphael Enrique Tiongco, M.Sc. for the free consultation regarding the framework of the overall meta-analysis. Moreover, the authors would like to extend their gratitude to the Angeles University Foundation for providing them with materials concerning genetic susceptibility of the interleukin-1 and as well as providing us with the EBSCOHost search engine that considerably aided us to complete the study.

